# Mutual relationships between head injury and conduct problems in children aged 9 months to 14 years in the UK Millennium Cohort Study

**DOI:** 10.1101/2022.01.09.22268970

**Authors:** Valerie Brandt, Charlotte Hall, Hedwig Eisenbarth, James Hall

## Abstract

**Background:** Research suggests a link between acquired head injury and signs of conduct disorder, with a majority of findings based on retrospective reports and comparison samples. The relationship between head injuries and conduct problems and how they may influence one another during development is currently unclear. This study aimed to investigate direct and indirect associations between head injury and conduct problems through to early adolescence.

**Methods:** Data from the UK Millennium Cohort Study was used to investigate the relationship between conduct problems as assessed by the Strengths and Difficulties Questionnaire and parent reported head injury over time, at ages 9 months, 3, 5, 7, 11 and 14 years, using a cross-lagged path analysis. This is data from 18,552 children, participating in a UK cohort study that is representative of the UK population. We included 7,041 (3,308 male) children, who had full information about head injuries and conduct problems at age 14.

**Results:** We found a mutual association between childhood head injuries and conduct problems but with distinct timings: Head injury between 5-7 years predicted greater chance of conduct problems at age 11 and 14 years, while greater conduct problems at 5 years predicted a significantly greater chance of a head injury at age 7-11 years.

**Conclusions:** These findings have important implications for the timing of preventive and ameliorative interventions. Prior to school entry, interventions aiming to reduce conduct problems would appear most effective at reducing likelihood of head injuries in future years. However, equivalent interventions targeting head injuries would be better timed either as children are entering formal primary education, or soon after they have entered.

## Introduction

Conduct problems encompass aggressive, destructive and deceptive behaviour, such as violating rules or social norms or breaking the law. If these problems are an ongoing pattern, conduct disorder can be diagnosed^1^. Conduct disorder is associated with risk taking behaviour^2,3^, probably due to impaired fear conditioning, hyporeactivity to incentives and impairments of the paralimbic system^4^. While research has shown that losing one’s temper and displays of mild aggression are common amongst healthy children^5^, aggressive behaviour that overcomes expected age-related and social norms in childhood has a high degree of persistency^6,7^. This is often followed by clinically significant problems arising in adolescence and adulthood ^8-11^, such as greater delinquency, violence, poor mental and physical health, and economic problems^11^. Potential causes of conduct problems in young people are diverse and include social, familial and neurodevelopmental risk factors^11^. One of these risk factors is head injury.

Head injury is the main cause of death and disability for children and adolescents in the UK ^12^. Up to 1.4 million individuals attend hospital for a head injury in England and Wales each year, and around 33% to 50% of those are children and adolescents younger than 15 years old^12^. Although head injuries and TBIs are not clearly distinct, partially because most individuals with a head injury will not receive a brain scan, a difference between mild head injuries and TBIs can be made based on clinical criteria. The national institute for health and care excellence (NICE) define head injuries as any trauma to the head excluding superficial facial injuries^12^, while it defines TBIs as head injuries resulting in structural damage and/or new onset or worsening disruption of brain function such as amnesia or aphasia^12^.

Some studies have found a relationship between head injuries and aggressive or delinquent behaviour. For instance, some clinical studies have shown increased aggressive behaviour following TBIs^13-17^, and TBIs have been shown to be more common in offenders than in non-offenders^18-20^. Two longitudinal studies showed that head injuries in adolescence predicted more interpersonal violence in early adulthood^21^, and violent behaviour one year later^22^. However, pre-existing conduct problems also have also been shown to be exacerbated by TBIs^23-29^ and it is therefore important to take into account pre-injury levels of aggressive behaviour when assessing the effects of TBIs or head injuries more generally.

Only 10.9% of TBIs that are reported to an emergency department are moderate or severe^30^. Mild head injuries without severe consequences (e.g. bumping one’s head without losing consciousness) are common in childhood^31^, and their often persistent and progressive long-term consequences are less well understood^32^. It is therefore largely unclear how head injuries and conduct problems may be related across development. Furthermore, the direction of the association between head injury and conduct problems is currently unclear, i.e. does a head injury lead to more conduct problems or do conduct problems increase the likelihood of head injuries? This is particularly true for children. Reflecting on the source of the current uncertainty in the literature, there are a number of methodological limitations including: a) frequent use of small and niche populations (clinical samples, moderate – severe TBI, criminal offending populations), which limits generalisability; b) reliance on retrospective, self-report data, which limits reliability and validity; and c) a use of cross-sectional designs, which do not allow a developmental interpretation of the relationship between head injuries and conduct problems.

Therefore, the current study uses a nationally representative UK cohort that were surveyed six times from age 9 months to 14 years between 2000-2015 to investigate the relationship between head injury and conduct problems in children over time. To account for other well-established risk factors for conduct problems, such as male gender and low family income^33^, these were included as covariates in this study. Based on previous research we hypothesised that conduct problems and incidence of head injury exacerbate each other over time.

## Methods

This study uses Millennium Cohort Study (MCS) data^34-38^, which follows the development of 19,517 children born between 2000-2002 in the UK^39^. Families were initially recruited via random sampling using all listed parents on the UK Child Benefit registers. The study is disproportionally stratified to allow areas of minority (e.g. ethnic minorities and disadvantaged areas) to be sufficiently signified, resulting in a nationally representative cohort. The MCS includes weight variables to allow for accurate representation of all population groups. Interviews with parents took place when the children’s age was 9 months (T1), 3 (T2), 5 (T3), 7 (T4), 11 (T5), and 14 years (T6). Parents gave written informed consent. This secondary data analysis was approved by the University of Southampton Ethics Committee.

### Participants

Only first-born children were included in this study, to minimise family related extraneous variables^40,41^. After matching data across all time points for first-borns (T1-6), the dataset included n = 18,552 participants. In addition, children were excluded if they had a diagnosis of Attention Deficit Hyperactivity Disorder (ADHD) or any indicators of epilepsy (e.g. seizures, Febrile fits and Fainting due to seizures) at any MCS data sweep, due to the increased risk of head injury associated with both diagnoses^42,43^. *N* = 497 had an ADHD diagnosis, n = 887 had an epilepsy diagnosis, n = 45 had both, overall excluded n = 1339. There are different possibilities to handle missing date. For our analyses, only participants with no missing data for the outcome variables at the latest data sweep of the MCS (T6) were included^44^, resulting in n = 7041 (n = 3308 male) participants ^45^.

### Measures

#### Conduct Problems

The Strengths and Difficulties Questionnaire (SDQ)^46-48^ has 25 items with a three-point rating scale (0 = not true, 1 = somewhat true, 3 = certainly true), and five subscales: hyperactivity / inattention, emotional difficulties, conduct problems, peer problems and the prosocial behaviour subscale, with good overall psychometric properties and good performance as screening instruments for a number of psychiatric diagnoses^49,50^. SDQ parent-rated data was collected from T2 (age 3). This study used the conduct problems subscale (5 items), with questions regarding aggressive behaviour, e.g. ‘*often fights with other children or bullies them*’. A higher score indicates a higher level of conduct problems (possible range: 0-10).

#### Head injury

At each data sweep parents reported if their child had experienced any accidents or injuries since the last data sweep. If yes, the frequency of accidents/ injuries was recorded and several injuries could be further defined: ‘*Thinking about the most severe (or only) accident or injury, what sort of accident or injury was it?*’ The responses were then coded into categories by interviewers (For the full list of injury categories see supplement 1). For the current study, a binary measure of Head Injury was constructed:

1. Bang to the head (coded 1) encompassing those coded with a ‘loss of consciousness/knocked out’, or ‘injury to the head without being knocked out’,
2. No head injury (coded 0) including no injuries and every injury other than head injury.

#### Covariates

Income and gender have been found to be associated with problematic behaviour^33^. Cohort member’s gender (n = 3308 male [47%]) and OECD equivalized household income (mean = 315, SD = 205, n = 6968) from T1 were therefore included in the path analysis. Males were coded 1, females 2.

#### Data analysis

Weights were used. Mplus 7.4^51^ was used to run a cross-lagged path analysis (Figure 1) to test reciprocal influences on head injury and conduct problems, with weighted least square means to account for missing data. Path analysis involves estimating direct, indirect, and total effects (which is the sum of the direct effect plus all indirect effects). The indirect effects can also be summed to produce a total indirect effect. For a classic primer on direct, indirect, and total effects see^52^. These effects are estimated as matter of course within the Mplus software.

**Figure 1:**
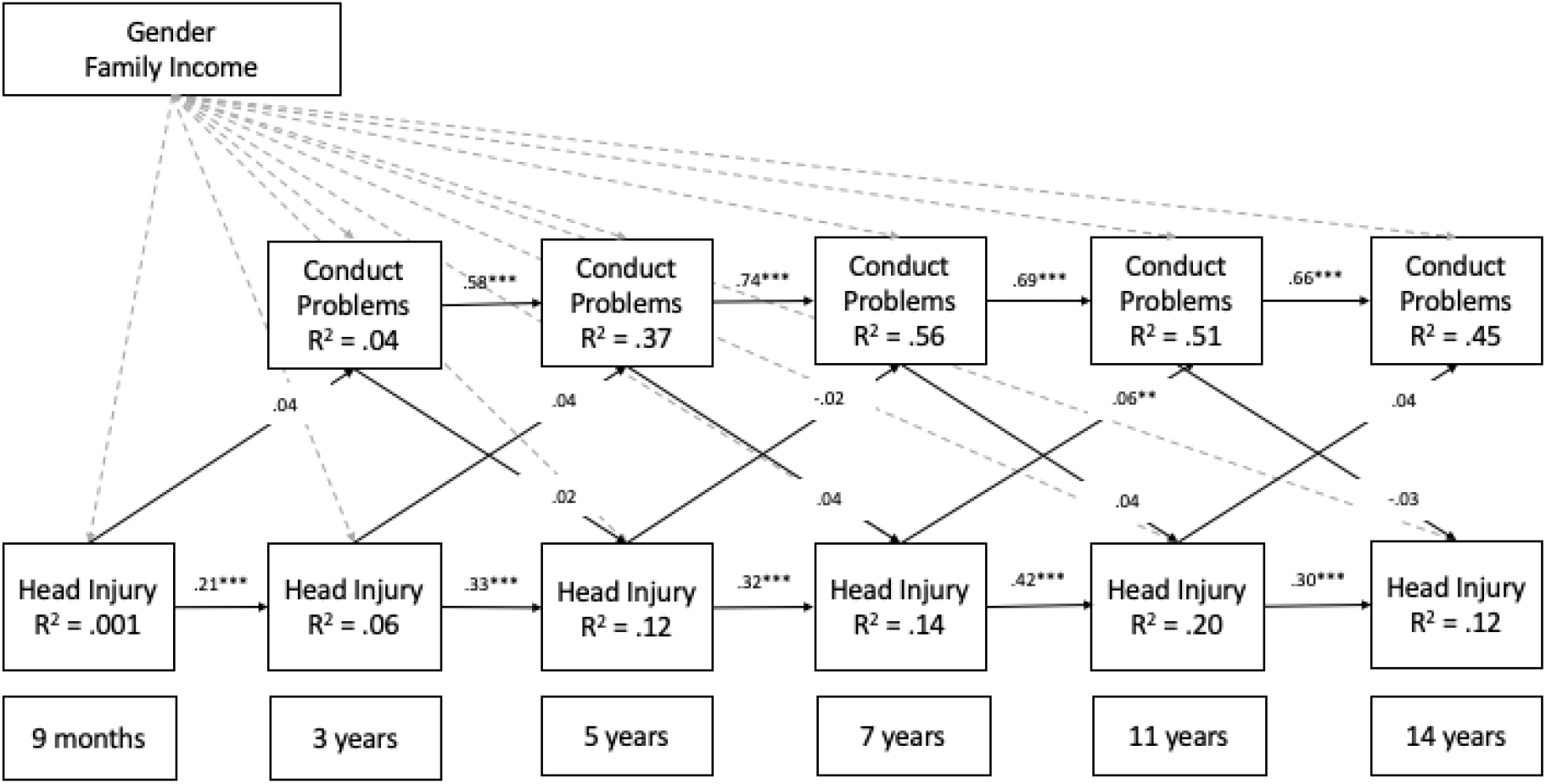
Cross-lagged path analysis of head injury (HI) and conduct problems (CP), over six time points aged 9 months – 14 years. Each arrow is accompanied by the appropriate standardised effect (Beta when CP is the dependent, Z when HI is the dependent). Standardized effects (same metrics) from the covariates can be found in Table 2. \**p* < .05 \*\**p* < .01 *** *p* < .001

Error terms were included for all indicators for consideration of extraneous factors. Satisfactory model fit was evaluated via the model fit indicators: Tucker-Lewis Index (TLI; satisfactory values ∼0.96), the Comparative Fit Index (CFI; satisfactory values ∼0.95), WRMR, and Root Mean Square Error of Approximation (RMSEA; satisfactory values <.06). Where the dependent variable is continuous (CP), standardised beta values with significance levels are reported. For the binary variable head injury, the standardised z-value (index of probit regression) is reported. Results are considered significant with α=.05.

### Data availability

Data are available through the Millennium Cohort Study (MCS) ^34-38^.

## Results

### Cross-lagged path analysis

Table 1 shows the descriptive data for number of head injuries and conduct disorder scores over time. The cross-lagged path analysis of head injury and conduct problems for UK children aged 9 months – 14 years in the Millennium Cohort Study showed acceptable model fit^**53**^ with an RMSEA of .04, CFI of .94, TLI of .88 and WRMR of 2.04 (χ^*2*^ _(36)_ = 526.19, *p* < .001).

**Table 1.**
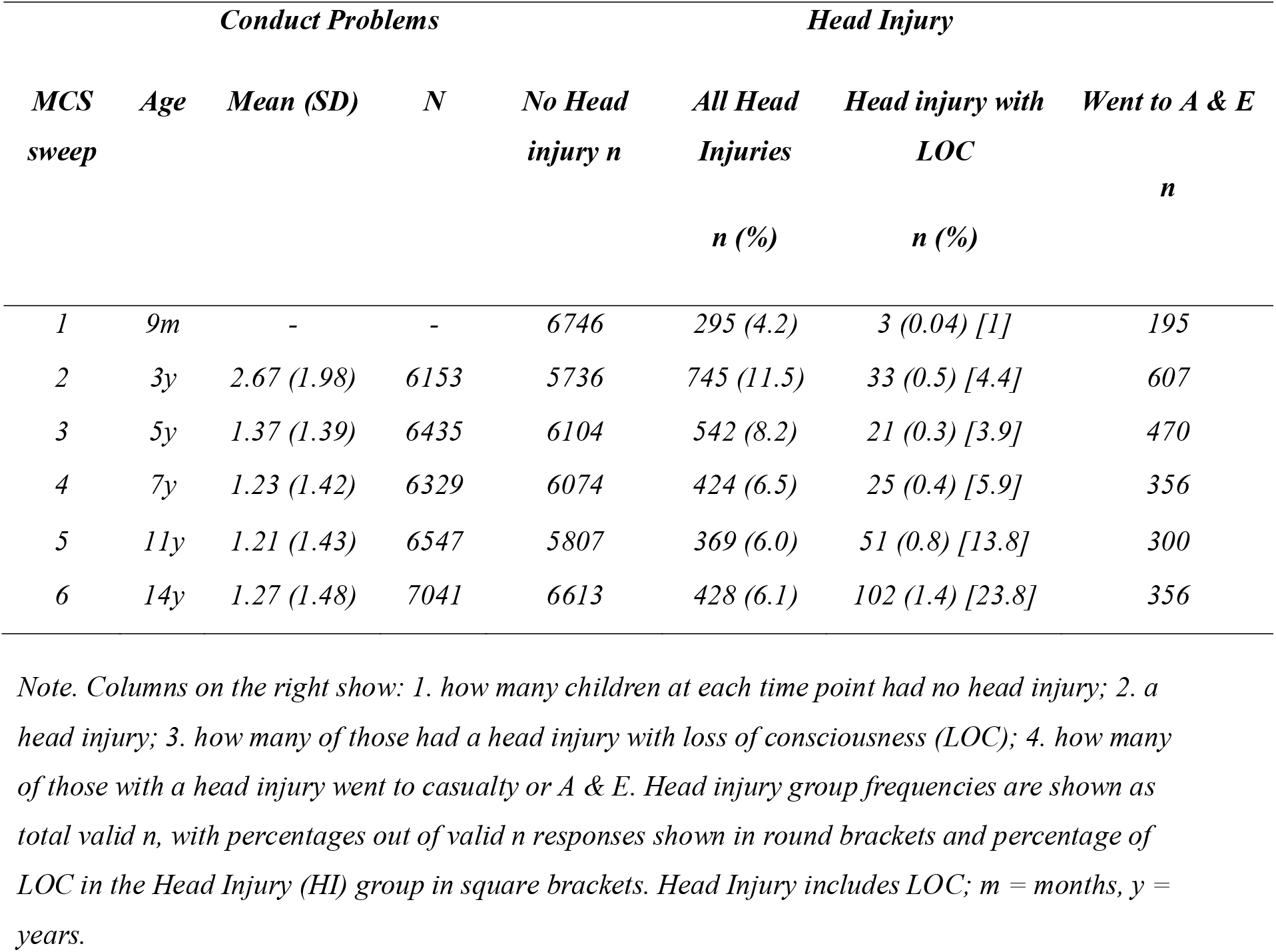
Descriptive Statistics for SDQ Conduct Problems and Head Injury

| *Conduct Problems* |  |  |  | *Head Injury* |  |  |  |
| --- | --- | --- | --- | --- | --- | --- | --- |
| *MCS sweep* | *Age* | *Mean (SD)* | *N* | *No Head injury n* | *All Head Injuries n (%)* | *Head injury with LOC n (%)* | *Went to A & E n* |
| 1 | 9m | - | - | 6746 | 295 (4.2) | 3 (0.04) [1] | 195 |
| 2 | 3y | 2.67 (1.98) | 6153 | 5736 | 745 (11.5) | 33 (0.5) [4.4] | 607 |
| 3 | 5y | 1.37 (1.39) | 6435 | 6104 | 542 (8.2) | 21 (0.3) [3.9] | 470 |
| 4 | 7y | 1.23 (1.42) | 6329 | 6074 | 424 (6.5) | 25 (0.4) [5.9] | 356 |
| 5 | 11y | 1.21 (1.43) | 6547 | 5807 | 369 (6.0) | 51 (0.8) [13.8] | 300 |
| 6 | 14y | 1.27 (1.48) | 7041 | 6613 | 428 (6.1) | 102 (1.4) [23.8] | 356 |

Note. Columns on the right show: 1. how many children at each time point had no head injury; 2. a head injury; 3. how many of those had a head injury with loss of consciousness (LOC); 4. how many of those with a head injury went to casualty or A & E. Head injury group frequencies are shown as total valid n, with percentages out of valid n responses shown in round brackets and percentage of LOC in the Head Injury (HI) group in square brackets. Head Injury includes LOC; m = months, y = years.

Considering direct effects linking variables at time T to time T+1: conduct problems at each time point significantly predicted conduct problems at the following time point as did head injury. For direct and indirect effects linking variables at time T to variables at time points later than T+1: head injury at age 7 had a significant direct effect on conduct problems at age 11 (Figure 1) and an indirect effect on conduct problems at age 14 (Table 3). Conduct problems at age 3 had a significant indirect effect on head injury at age 11 (Table 3) but there were no significant direct effects of conduct problems on head injury (Figure 1, Table 3).

**Table 2:** Direct (standardised) effects of gender and family income on head injury (HI) and conduct problems (CP) (statistically significant results are marked in bold)

|  | **B** | **S.E.** | **P** |
| --- | --- | --- | --- |
| **Conduct Problems 2** |  |  |  |
| Income | - 0.20 | 0.01 | < **0.001** |
| Females | - 0.03 | 0.02 | **0.023** |
| **Conduct Problems 3** |  |  |  |
| Income | - 0.09 | 0.02 | < **0.001** |
| Females | - 0.04 | 0.01 | **0.004** |
| **Conduct Problems 4** |  |  |  |
| Income | - 0.05 | 0.02 | **0.001** |
| Females | - 0.03 | 0.01 | **0.025** |
| **Conduct Problems 5** |  |  |  |
| Income | - 0.07 | 0.02 | < **0.001** |
| Females | - 0.01 | 0.01 | 0.530 |
| **Conduct Problems 6** |  |  |  |
| Income | -0.04 | 0.01 | **0.001** |
| Females | 0.04 | 0.01 | **0.002** |
|  | **Z** | **S.E.** | **P** |
| **Head Injury 1** |  |  |  |
| Income | - 0.03 | 0.03 | 0.404 |
| Females | - 0.01 | 0.03 | 0.711 |
| **Head Injury 2** |  |  |  |
| Income | - 0.02 | 0.03 | 0.335 |
| Females | - 0.12 | 0.02 | < **0.001** |
| **Head Injury 3** |  |  |  |
| Income | - 0.04 | 0.03 | 0.179 |
| Females | - 0.08 | 0.03 | **0.003** |
| **Head Injury 4** |  |  |  |
| Income | - 0.07 | 0.03 | **0.038** |
| Females | - 0.12 | 0.03 | < **0.001** |
| **Head Injury 5** |  |  |  |
| Income | - 0.02 | 0.03 | 0.519 |
| Females | - 0.06 | 0.03 | **0.042** |
| **Head Injury 6** |  |  |  |
| Income | - 0.01 | 0.03 | 0.719 |
| Females | - 0.14 | 0.03 | < **0.001** |

1 *Head injury at waves 1-6, Conduct problems (CP) at waves 2-6, see ages for each wave in Table 1.*

**Table 3.**
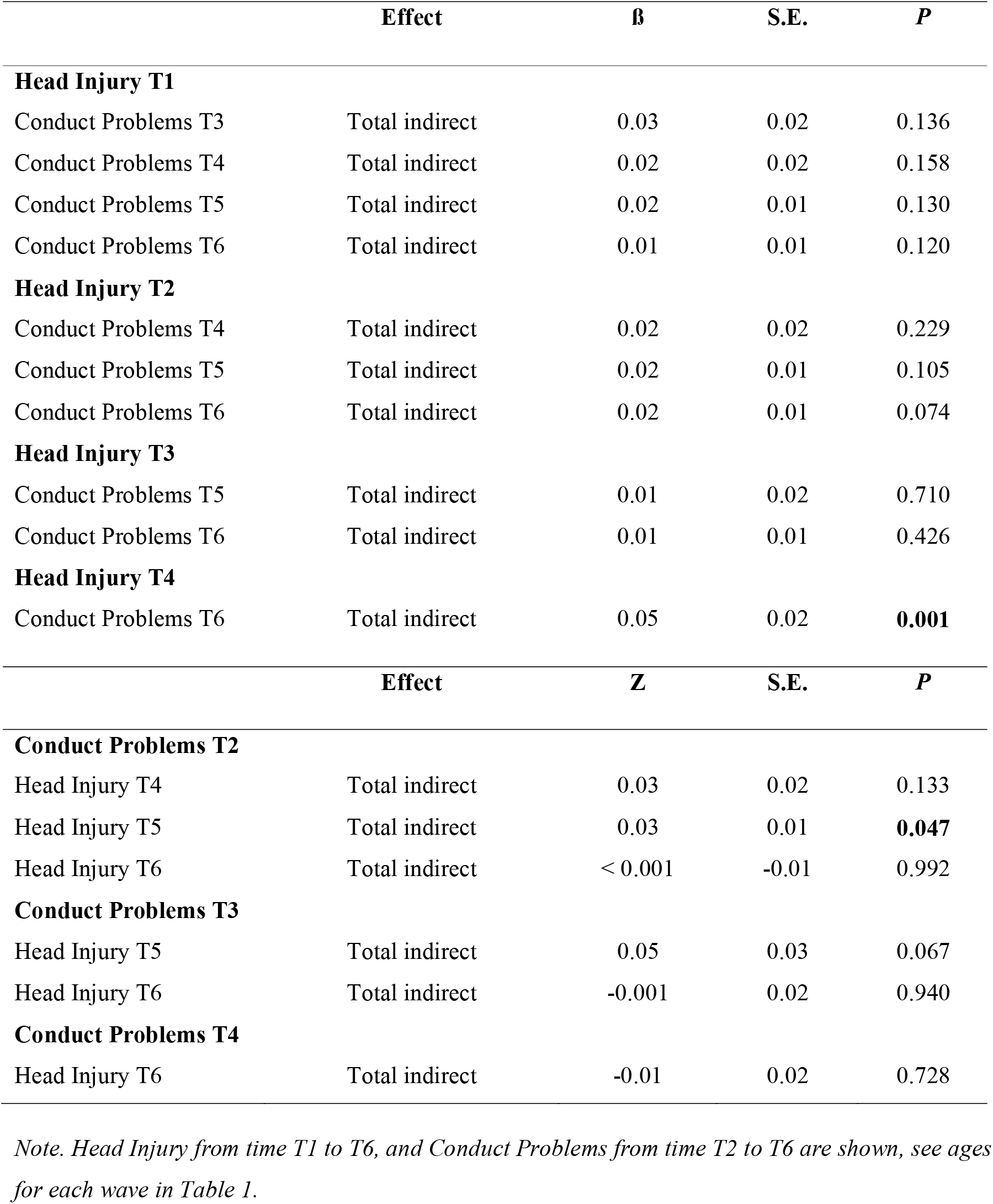
Total indirect effects for head injury on conduct problems and for conduct problems on head injury

### Covariate effects

Lower income was significantly related to higher conduct problem at all ages (3 – 14) but to higher likelihood for head injury only at age 11 (Table 2). Male gender was significantly related to higher conduct problems scores age 3-7 and to a higher likelihood for obtaining a head injury at all ages from 3-14 years. Surprisingly, females showed greater average parent-reported conduct problems at age 14 than males.

## Discussion

This study aimed to characterise the relationship between conduct problems and head injuries between age 9 months to 14 years in a sample of children representative of the general population in the UK. The results show three ways in which head injuries and conduct problems are associated across development. Most importantly though, head injuries at most ages did not lead to a direct increase in conduct problems or vice versa. Only head injury between 5-7 years was a significant risk factor for conduct problems (at age 11 and 14). Further, only greater conduct problems between 1-3 years was a risk factor for head injury (at age 11).

Setting these results in context, previous studies in community samples have found increased problematic behaviour in adolescents following or associated with a head injury^22,54^. However, the initial assessment by Buckley & Chapman (2017) included adolescents with a mean age of 13.45, whereas participants in the last wave of the current study were 14 years old. Ilie et al. (2014) investigated cross-sectional data and only investigated head injuries with loss of consciousness, while the majority of head injuries in general, and in this sample, did not result in loss of consciousness. Severity of head injury (e.g. loss of consciousness, amnesia) may play a role with regard to conduct problems but results have been mixed. A small study found that children with mild TBIs did not differ from children with orthopaedic injuries and both were significantly less aggressive than children with severe TBIs^55^, while other studies found that TBIs of all severity were associated with more conduct problems^17,56^. Catroppa and colleagues (2008) showed that injury severity significantly predicted post-injury behavioural problems in 2-6 year olds, however, behaviour pre-injury was the strongest predictor and should be controlled for^57^. While only a minority of head injuries in this study resulted in loss of consciousness, injuries were significant enough that most children were taken to hospital. The data are representative of the UK population and therefore provide a clearer picture of the relationship between head injury and conduct problems, without focusing on severe cases. The findings are important because head injuries are common but are mild in the majority of cases^31^. It would be interesting to explore the difference between head injuries with and without a loss of consciousness. However, the number of children with a severe head injury was too small for a complex model, particularly at T1 (Table 1).

The exact relationship between age at injury and conduct problems has also remained unclear, with some contradictory findings^25,58,59^. A twin study showed that head injuries early in childhood were associated with a delayed decline in impulsivity and greater increase in reactive aggressiveness by adolescence^60^. However, head injury was assessed retrospectively. Our results indicate that age at injury plays a role. While head injuries do not lead to higher levels of conduct problems at most ages, it appears that they can, if occurring between age 5-7. Our results also showed that there was an indirect effect of conduct problems at age 3 on likelihood of head injury at age 11. This is roughly in line with a previous study showing that aggressive behaviour predicts rate of any accidental injuries in 5-10 year olds^61^. Previous cross-sectional studies regarding the link between head injury and aggressive behaviour^21,22,28,54,59,62-64^ were unable to determine whether head injury leads to aggression, or if aggression leads to head injury^65^. Our results suggest that both can be the case.

The results have important implications for the timing of preventive and ameliorative interventions. Interventions aiming to reduce conduct problems might be most effectively tailored to children before they enter school, to reduce likelihood of subsequent head injury. Interventions targeting the prevention of head injury in order to reduce subsequent conduct problems might be most effective when children enter formal primary education, or in the early period of primary education. The education system could facilitate interventions in early primary school. For instance, accident prevention, particularly in boys, might be an effective strategy to reduce head injuries. It would be useful to identify where most head injuries occur (school or home setting, sports, etc.). Interventions identifying and treating conduct problems prior to school entry may be more difficult and involve parent training or screening by the GP. Existing parent trainings for conduct problems in children^66^ should include head injury prevention aspects.

Conduct problems decreased from age 3 over time. Furthermore, conduct problems significantly predicted conduct problems at each following time point. This finding is in line with previous research, showing that physical aggression tends to decrease in early childhood in the majority of children^67,68^ and that children typically follow stable trajectories^67,69^; for review see^70^. Interestingly, females had greater average parent-reported conduct problems at age 14 than boys (while controlling for income). Previous research in the MCS dataset has pointed to a group of children who have a school-onset trajectory of conduct problems and this was more common in girls than in boys^71^. Moreover, while physical aggression is more common in boys, other symptoms of conduct problems, such as lying and cheating, can be more prevalent in girls^72^. Future research should address the question which factors change girls’ conduct problem trajectories during adolescence.

In line with previous research^73^, our results also revealed that those who have experienced one head injury, are likely to experience another. Several risk factors may play a role in repeated head injuries. Some people may be more prone to having accidents than others^74^. Particularly head injuries are often followed by a range of, sometimes subtle, physical, cognitive, and affective changes^75^. As our data were based on general population interviews rather than clinical data, parental factors, such as a higher likelihood to seek medical treatment by some parents, cannot explain this stable effect. Another risk factor for head injury is physical abuse^76,77^, which is also a risk factor for conduct problems^78^. If head injuries and conduct problems are not directly associated at most ages, previous findings of associations between the two variables may be better explained by other common variables that influence both head injury and conduct problems, such as environmental factors. We did not determine specific risk factors for conduct problems and head injury other than gender and income^30,79^. Future studies could include further risk factors for conduct problems, such as other types of adverse life events, in order to further dissociate specific impact of brain injury from environmental factors.

Males were more likely than females to experience a head injury after the age of 9 months. This is in line with previous research, showing that being male and having a lower SES increases the risk for experiencing a head injury^30^. Our data confirms that these factors also increase the risk of displaying conduct problems^33^ though income only played a minor role in the risk for head injury. Future studies should investigate risk factors for repeated head injuries and their interactions in more detail.

Strengths of this study are the large number of children, its longitudinal nature and its naturalistic setting, i.e. all head injuries (with and without loss of consciousness) were compared to a control group of all others (without injury and all injuries other than head injury). We can therefore draw conclusions based on the general population, not on a clinical sample. Furthermore, head injuries and conduct problems were assessed at each time point and should not be as biased by memory as other retrospective data. Limitations include that head injuries were broadly defined, only first-borns were included, and that we were unable to differentiate between severe and less severe head injuries. Moreover, we used a fairly simple model, only controlling for the main risk factors for conduct problems and head injuries, i.e. gender and family income. Future studies should take into account further risk factors.

## Conclusions

Based on longitudinal data from the general population in the UK, we find that head injuries between 5-7 years are associated with increased conduct problems at ages 11 and 14 and that conduct problems between 3-5 years are associated with an increased risk for head injuries at age 11 years. These findings show which time windows during development might be most important for applying interventions that target accident prevention and conduct problems with regard to their mutual association over time.

## Data Availability

All data produced are available online at: https://ukdataservice.ac.uk

